# Circulating CD34^+^ Progenitor Cells are Predictive of All-Cause and Cardiovascular Mortality and are Influenced by Physical Activity.

**DOI:** 10.1101/2020.06.24.20139220

**Authors:** David Muggeridge, Jennifer Dodd, Mark D. Ross

## Abstract

**Background:** Circulating progenitor cells (CPCs) play an important role in vascular repair and may influence cardiovascular (CV) health and longevity. Exercise is known to modulate these cells via mobilization from the bone marrow. The primary aims of this study were to evaluate the association of CPCs with mortality and explore the association between physical activity (PA) and CPCs.

**Design:** We studied 1,751 individuals from the Framingham Offspring cohort (66 ± 9 years [40-92 years], 54% female). CPCs (CD34^+,^ CD34^+^CD133^+^, CD34^+^CD133^+^KDR^+^) were measured in participants using flow cytometry. Multivariable cox regression analyses were performed to investigate relationship of CPCs with future CV event, CV mortality and all-cause mortality. Multivariate regression analyses were performed to determine the relationship between self-reported PA and CPC counts.

**Results:** Following adjustment for standard risk factors, there was an inverse association between CD34^+^ CPCs and all-cause mortality (hazard ratio (HR) per unit increase in CD34^+^, 0.79; 95% CI 0.64 – 0.98, *P*=0.036). CD34^+^CD133^+^ CPCs were inversely associated with CV mortality (HR 0.63, 95% CI 0.44 – 0.91, *P*=0.013). Associations of CD34^+^ and CD34^+^CD133^+^ with mortality were strongest in participants with pre-existing CVD. PA was associated with CD34^+^ CPCs only in CVD participants. This relationship was maintained after adjustment for confounding variables.

**Conclusions:** Higher number of CD34^+^ and CD34^+^ CD133^+^ CPCs were inversely associated with all-cause and CV mortality. These associations were strongest in participants already diagnosed with CVD. PA is independently associated with CD34^+^ CPCs in individuals with CVD only, suggestive of greater benefit for this population group.

## Introduction

Circulating progenitor cells (CPC) are a heterogenous group of cells which have tissue regenerative potential. A number of studies have shown that CD34^+^ CPCs and several subsets of CD34^+^ cells (such as CD34^+^CD133^+^/KDR^+^) can participate in vascular repair and growth ^1, 2^, and may be associated with vascular endothelial function^3^. Therefore these cells may reflect vascular integrity and have been used as vascular biomarkers of repair ^4^.

Low number of these CPCs are associated with vascular dysfunction ^3, 5^ and subsequent greater cardiovascular (CV) risk ^6^. Observational studies have shown that individuals with cardiovascular disease (CVD) exhibit lower number and angiogenic function of these CPCs ^7^, reflecting reduced vascular repair capacity. Studies have demonstrated that in individuals hospitalized with heart failure ^8^, or with acute coronary syndromes ^9^, low number of CD34^+^ CPCs predict earlier mortality in these patients compared to patients with high numbers of CD34^+^ CPCs, which suggests impaired vascular repair capacity in those with higher mortality risk. Whilst there are no studies that have investigated the role of CD34^+^ CPCs and associated subsets in predicting clinical endpoints in a heterogenous human population, there is evidence to suggest that these CPCs are reflective of subclinical atherosclerotic risk in an apparently healthy population ^7^.

Lifestyle behaviors can significantly affect CV health. Smoking, physical inactivity and obesity are associated with perturbed vascular health, leading to greater risk of mortality. Physical activity, known for its effect on improving vascular function ^10^ may do so in part via modulating CPC content and/or function. Studies investigating acute ^11^ and chronic exercise training ^12^ have demonstrated that progenitor cells can be mobilized into peripheral blood compartment in humans, where they can exert their vasoreparative functions. However, the efficacy of exercise training to promote progenitor cell number has been argued, with recent evidence demonstrating little or no change in CPC number in humans after exercise training ^4^. As yet, there is no evidence from large cohorts investigating the association between physical activity and CPCs.

The primary aim of this study was to investigate the prognostic potential of CD34^+^ CPCs on all-cause and CV mortality. Furthermore, we investigated the relationship between self-reported physical activity on CPCs in a large cohort. We hypothesized that circulating CD34^+^ CPCs and subpopulations would predict mortality, and that these cells are associated with self-reported physical activity levels.

## Methods

### Study Sample

The Framingham Heart Study (FHS) is a longitudinal community-based cohort set up in 1948 under the direction of the National Heart, Lung, and Blood Institute (NHLBI) aimed to determine factors that contribute to the onset and progression of cardiovascular disease (CVD)^13^. Subsequently, an Offspring cohort was included from 1971^14^. Participants (n=3,002) in the Framingham Offspring cohort who attended the 8^th^ examination cycle (2004-2008) were eligible for our investigation, with n=1,751 included in the study due to availability of key data. Participant characteristics are shown in **Table 1**.

**Table 1.** Participant Characteristics.

|  | All<br>(n=1751) | CVD-free<br>(n=1467, 84%) | CVD<br>(n=284, 16%) | p-value |
| --- | --- | --- | --- | --- |
| Age (years) | 66 ± 9<br>[40-92] | 65 ± 9<br>[40-90] | 72 ± 9<br>[51-92] | <0.001 |
| Female (n, %) | 940, 53.7% | 822, 56% | 118, 41.5% | - |
| BMI<br>(kg·m<sup>2</sup>) | 28.4 ± 5.3<br>[13.8-54.2] | 28.2 ± 5.3<br>[13.8-54.2] | 29.2 ± 5.2<br>[18.4-45.1] | 0.007 |
| Systolic Blood<br>Pressure (mmHg) | 129 ± 18<br>[76-204] | 129 ± 18<br>[76-204] | 131 ± 19<br>[90-198] | 0.070 |
| Diastolic Blood<br>Pressure (mmHg) | 74 ± 11<br>[34-122] | 75 ± 10<br>[34-122] | 70 ± 11<br>[40-108] | <0.001*** |
| Fasting Glucose<br>(mg/dL) | 107.0 ± 24.1<br>[36-327] | 106 ± 23<br>[58-327] | 113 ± 28<br>[36-292] | <0.001*** |
| Total Cholesterol<br>(mg/dL) | 186.0 ± 37.5<br>[71-322] | 190 ± 35.6<br>[96-322] | 164 ± 39.4<br>[71-289] | <0.001*** |
| HDL-Cholesterol<br>(mg/dL) | 57.4 ± 18.2<br>[21-147] | 58.8 ± 18.3<br>[21-152] | 50.3 ± 15.8<br>[23-130] | <0.001*** |
| Triglycerides<br>(mg/dL) | 118.0 ± 70.6<br>[30-976] | 115 ± 67.2<br>[30-976] | 133 ± 84.4<br>[40-583] | <0.001 |
| Smokers (n,%) | 206, 11.8% | 170, 12% | 36, 13% |  |
| Hypertensive (n, %) | 989, 56.5% | 831, 57% | 158, 56% | - |
| CD34+ CPCs<br>(% MNCs) | 0.0873 ± 0.0492<br>[0.011-0.490] | 0.0875 ± 0.0483<br>[0.0110-0.490] | 0.0860 ± 0.0538<br>[0.0160-0.370] | 0.201 |
| CD34+CD133+ CPCs<br>(% MNCs) | 0.0402 ± 0.0366<br>[0.0020-0.6090] | 0.0407 ± 0.0380<br>[0.0020-0.6090] | 0.0375 ± 0.0286<br>[0.0040-0.2420] | 0.290 |
| CD34+CD133+KDR+<br>(% MNCs) | 0.0040 ± 0.0037<br>[0.0001-0.0470] | 0.0040 ± 0.0037<br>[0.0001-0.0470] | 0.0041 ± 0.0039<br>[0.0002-0.0261] | 0.648 |
| ECFC (number of<br>colonies) | 43 ± 31<br>[0-196] | 43 ± 31<br>[0-196] | 42 ± 32<br>[0-178] | 0.470 |

Data are mean ± SD [range]. ECFC- Endothelial Colony Forming Cells (in 1631 participants). \* $p < 0.05$ , \*\*\* $p < 0.001$ .

This study complies with the Declaration of Helsinki. Ethical approval for all data collection and research purposes was granted by Boston University Medical Centre, and consent was obtained for the collection and use of the data available for secondary investigators. Edinburgh Napier University Research Ethics and Integrity Committee approved of the use of the secondary dataset for the purposes of the study.

### Clinical Assessment

All participants underwent a clinical and risk factor assessment including assessment of blood pressure, height and body mass. Fasting blood samples were drawn for quantification of glucose, glycated hemoglobin (HbA1c), total cholesterol, and high density lipoprotein cholesterol (HDL-C), and triglycerides.

### Quantification of Circulating Progenitor Cells

Blood samples were collected from participants in the fasted state to quantify CPC counts. Blood samples were centrifuged and the peripheral blood mononuclear cells (PBMCs) were isolated for cell phenotyping as previously described^15^. PBMCs were stained with anti-CD34 FITC, anti-CD133 APC and anti-KDR-PE antibodies (all BD Biosciences). CD34^+^ cells were gated for subsequent expression of CD133 and finally KDR. Total progenitor cells are defined as CD34^+^ cells, and EPCs are defined as CD34^+^CD133^+^ and CD34^+^CD133^+^KDR^+^ cells. Analysis of flow cytometry files were performed using FlowJo analysis software (Treeestar, Inc.) and reviewed by investigators blinded to the identity of the participants.

### Endothelial Cell Colony Forming Cells (ECFC)

In 1653 participants, PBMCs were also used to assess endothelial cell colony forming cells (ECFC). PBMCs were cultured on fibronectin-coated tissue culture plates (BD Biosciences) and cultured for 7 days. After 7 days of culture, the number of colonies in each well were counted by a single blinded individual. ECFC number was reported as average number of colonies per well up to 12 wells.

### Mortality and Event Incidence

Follow up was conducted for primary end points of all-cause and cardiovascular death. Cause of death was determined through medical history, review of medical records, death certificate, interview of next of kin, and review of the National Death Index. Cardiovascular death was defined as death attributed to ischemic cause (fatal myocardial infarction, stroke). Cardiovascular event risk was only assessed in individuals with no pre-existing CVD or CV event occurring before exam 8 (n = 1467). CV event or incident CVD was assessed using the standard Framingham Heart Study criteria and included the following: new-onset angina, fatal and non-fatal MI or stroke, heart failure or intermittent claudication.

### Self-Reported Physical Activity Levels

Self-report sleep, sitting time, slight, moderate and heavy activity were determined using a physical activity questionnaire employed by the Framingham Heart Study. The numbers of hours of certain activity per week were collected. A composite score was calculated (physical activity index; PAI), for each participant by weighting a 24hr activity recall. Participants were asked to report the number of hours in a typical day spent sleeping (weighting factor [WF] = 1) and in sedentary (WF = 1.1), slight (WF = 1.5), moderate (WF = 2.4), and heavy activities (WF = 5) ^16^. PAI was subsequently calculated by adding the products of the hours spent at each activity domain and their weighting factor based on the oxygen requirements for said activity 17.

### Statistical Analysis

Continuous variables were assessed for normality by assessing histograms and Q-Q plots. Data for CD34^+^, CD34^+^CD133^+^KDR^+^, CD34^+^CD133^+^ and PAI were natural log transformed and EFCFs square root Appropriate data transformations were applied where relevant prior to further analysis. All participants were categorized into tertiles for each CPC measure for event and mortality risk analyses using Kaplan-Meier curve and log-rank analyses. Subsequent Cox proportional hazards regression analyses were performed, utilizing transformed continuous data for CPC’s. Cox proportional hazards regressions were performed unadjusted and adjusted for age, sex, BMI, PAI, CVD and diabetes status, smoking status. To investigate the effects of CVD status, the data set was split and analyses repeated for those free of CVD at exam 8 (n = 1467) and those with a CVD diagnosis prior to exam 8 (n = 284). Proportional hazards assumptions for each of the Cox models were evaluated by plots of Schoenfeld residuals.

To assess the influence of physical activity on CPC counts, linear regression analyses were performed to assess the relationship between CPC counts and PAI. A subset of physical activity, moderate + heavy activity time, was also investigated. Unadjusted and adjusted analyses are displayed. Data analyses were carried out using RStudio Team (2019, RStudio: Integrated Development for R. RStudio, Inc, Boston, MA: http://www.rstudio.com/). *P*-values of <0.05 were considered statistically significant.

## Results

### Relationship between CPC Counts and Adverse Events

#### All-Cause Mortality

Kaplan Meier curves based on tertiles of CPC counts and all-cause mortality are shown in **Figure 1 A-D**. In unadjusted Cox proportional hazard models, increases in CD34^+^ and CD34^+^CD133^+^ CPCs were significantly associated with a decrease risk of death (*P* <0.001, *P =0*.*001*; **Table 2**). Following adjustment, increases in CD34^+^ remained significantly associated with a decrease risk of death (*P =* 0.036). Whilst there was a trend for CD34^+^CD133^+^ on all-cause mortality this did not reach statistical significance (*P =* 0.07). No significant associations were observed for all-cause mortality for CD34^+^CD133^+^KDR^+^ EPCs or ECFC (all *P* > 0.05; **Table 2**).

**Table 2.**
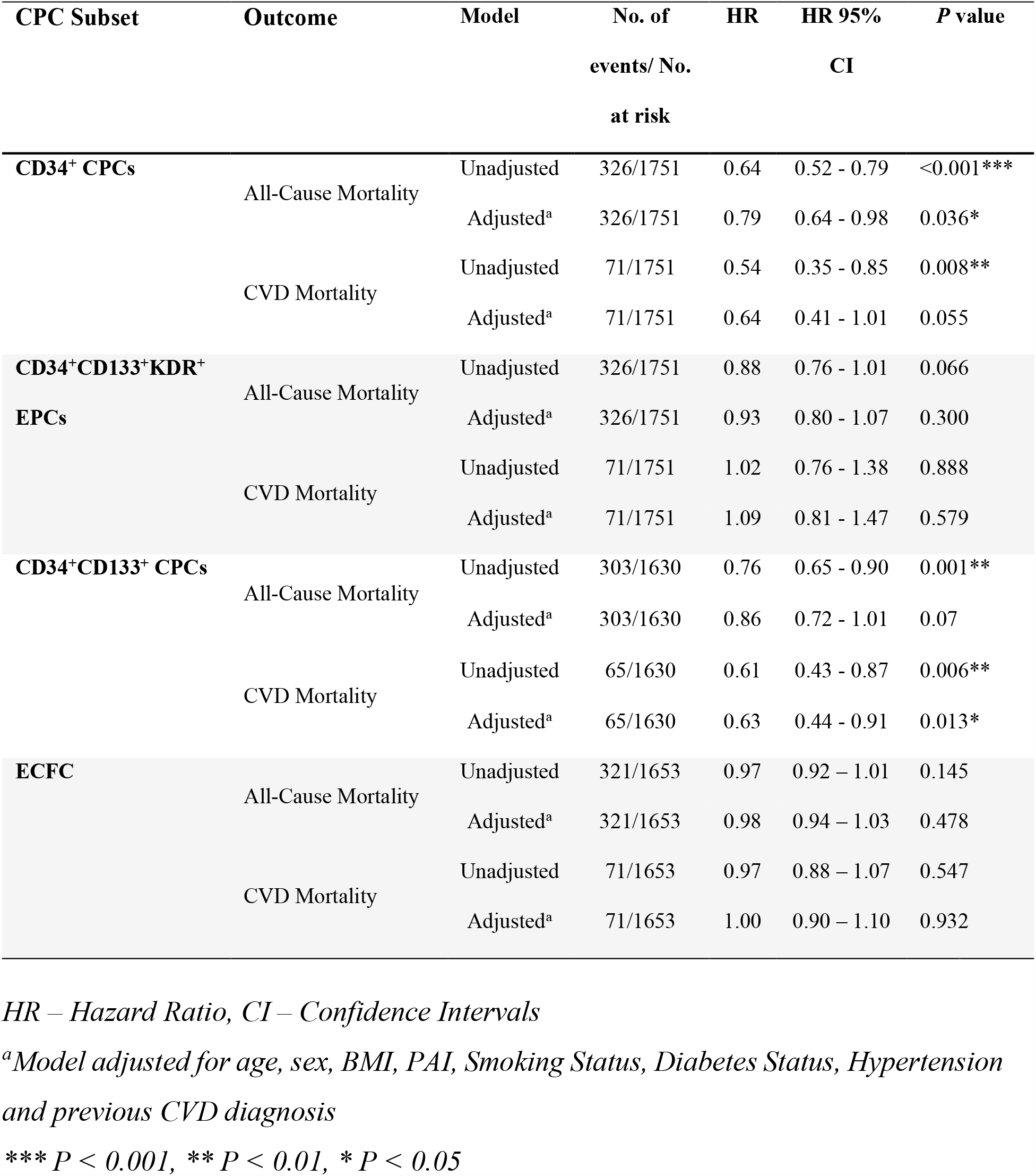
CPC Counts and risk of death for all participants.

| CPC Subset | Outcome | Model | No. of<br>events/ No.<br>at risk | HR | HR 95%<br>CI | P value |
| --- | --- | --- | --- | --- | --- | --- |
| **CD34<sup>+</sup> CPCs** | All-Cause Mortality | Unadjusted | 326/1751 | 0.64 | 0.52 - 0.79 | <0.001*** |
|  |  | Adjusted<sup>a</sup> | 326/1751 | 0.79 | 0.64 - 0.98 | 0.036* |
|  | CVD Mortality | Unadjusted | 71/1751 | 0.54 | 0.35 - 0.85 | 0.008** |
|  |  | Adjusted<sup>a</sup> | 71/1751 | 0.64 | 0.41 - 1.01 | 0.055 |
| **CD34<sup>+</sup>CD133<sup>+</sup>KDR<sup>+</sup>**<br>**EPCs** | All-Cause Mortality | Unadjusted | 326/1751 | 0.88 | 0.76 - 1.01 | 0.066 |
|  |  | Adjusted<sup>a</sup> | 326/1751 | 0.93 | 0.80 - 1.07 | 0.300 |
|  | CVD Mortality | Unadjusted | 71/1751 | 1.02 | 0.76 - 1.38 | 0.888 |
|  |  | Adjusted<sup>a</sup> | 71/1751 | 1.09 | 0.81 - 1.47 | 0.579 |
| **CD34<sup>+</sup>CD133<sup>+</sup> CPCs** | All-Cause Mortality | Unadjusted | 303/1630 | 0.76 | 0.65 - 0.90 | 0.001** |
|  |  | Adjusted<sup>a</sup> | 303/1630 | 0.86 | 0.72 - 1.01 | 0.07 |
|  | CVD Mortality | Unadjusted | 65/1630 | 0.61 | 0.43 - 0.87 | 0.006** |
|  |  | Adjusted<sup>a</sup> | 65/1630 | 0.63 | 0.44 - 0.91 | 0.013* |
| **ECFC** | All-Cause Mortality | Unadjusted | 321/1653 | 0.97 | 0.92 – 1.01 | 0.145 |
|  |  | Adjusted<sup>a</sup> | 321/1653 | 0.98 | 0.94 – 1.03 | 0.478 |
|  | CVD Mortality | Unadjusted | 71/1653 | 0.97 | 0.88 – 1.07 | 0.547 |
|  |  | Adjusted<sup>a</sup> | 71/1653 | 1.00 | 0.90 – 1.10 | 0.932 |

*HR – Hazard Ratio, CI – Confidence Intervals*
*<sup>a</sup>Model adjusted for age, sex, BMI, PAI, Smoking Status, Diabetes Status, Hypertension*
*and previous CVD diagnosis*
*\*\*\* P < 0.001, \*\* P < 0.01, \* P < 0.05*

**Figure 1.**
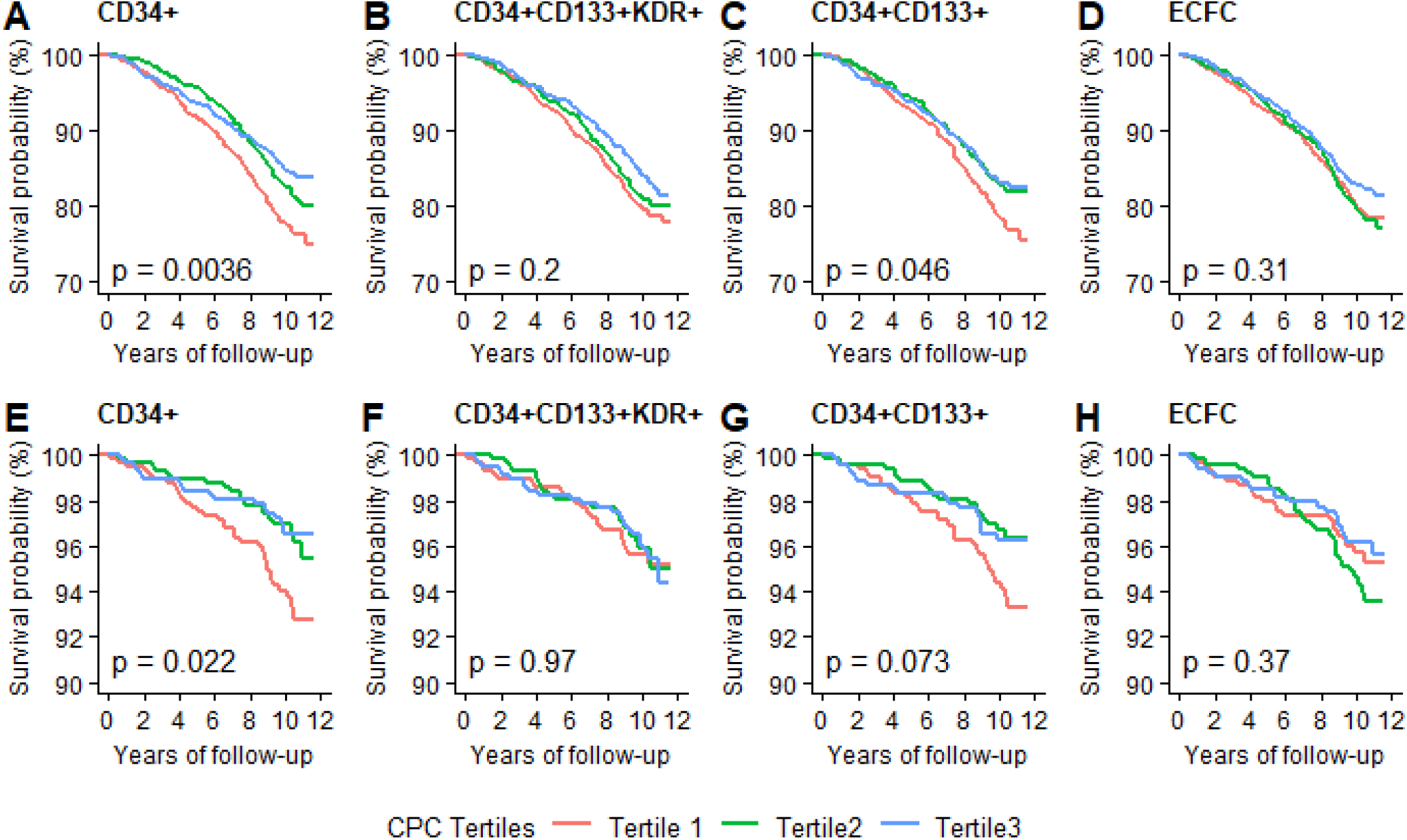
Kaplan Meier Survival Curve’s for the relationship between CPC tertile group and all-cause mortality (A, B, C, D) and cardiovascular mortality (E, F, G, H) (Tertile 1 = Low count, Tertile 2 = Moderate count, Tertile 3 = High count). Statistical significance was set at p <0.05.

#### Cardiovascular Mortality

Kaplan Meier curves based on tertiles of CPC counts and cardiovascular mortality are shown in **Figure 1 E-H**. In unadjusted Cox proportional hazard models, increases in CD34^+^ and CD34^+^CD133^+^ CPCs were significantly associated with a decrease risk of cardiovascular death (*P =* 0.008, *P =* 0.006; **Table 2**). Following adjustment, CD34^+^CD133^+^ CPCs were significantly associated with a decrease risk of cardiovascular death (*P* = 0.013). Whilst there was a trend for CD34^+^ on CVD mortality this did not reach statistical significance (*P* = 0.055). No other significant associations were observed for CVD mortality (all *P* > 0.05; **Table 2**).

### Relationship between CPC Counts and Adverse Events-Influence of CVD Status

#### All-Cause Mortality

Kaplan Meier curves based on tertiles of CPC counts and all-cause mortality for those free of CVD and those with CVD at exam 8 are shown in **Supplementary Figure 1**. Unadjusted and adjusted Cox proportional hazard models for CPC counts are displayed in **Table 3**. Following adjustment, increases in CD34^+^ and CD34^+^CD133^+^ CPCs were significantly associated with a decrease risk of death in those with CVD at exam 8 (*P =* 0.032, *P =* 0.003). No other significant associations were observed for all-cause mortality (all *P* > 0.05; **Table 3**).

**Table 3.** CPC Counts and risk of death for all participants split by CVD diagnosis at exam 8.

| CPC Subset | Outcome | Model | CVD Free at Exam 8 |  |  |  | CVD Diagnosis by Exam 8 |  |  |  |
| --- | --- | --- | --- | --- | --- | --- | --- | --- | --- | --- |
|  |  |  | No. of events/ No. | HR | HR 95% CI | P value | No. of events/ | HR | HR 95% CI | P value |
|  |  |  | at risk |  |  |  | No. at risk |  |  |  |
| **CD34<sup>+</sup> CPCs** | All-Cause Mortality | Unadjusted | 211/1467 | 0.70 | 0.54 - 0.91 | 0.008** | 115/284 | 0.57 | 0.41 - 0.81 | 0.002** |
|  |  | Adjusted<sup>a</sup> | 211/1467 | 0.89 | 0.67 - 1.18 | 0.424 | 115/284 | 0.68 | 0.48 - 0.97 | 0.032* |
|  | CVD Mortality | Unadjusted | 34/1467 | 0.65 | 0.34 - 1.26 | 0.203 | 37/284 | 0.49 | 0.26 - 0.91 | 0.023* |
|  |  | Adjusted<sup>a</sup> | 34/1467 | 0.84 | 0.41 - 1.70 | 0.619 | 37/284 | 0.53 | 0.28 - 0.99 | 0.048* |
| **CD34<sup>+</sup>CD133<sup>+</sup>KDR<sup>+</sup> EPCs** | All-Cause Mortality | Unadjusted | 211/1467 | 0.90 | 0.76 - 1.08 | 0.258 | 115/284 | 0.85 | 0.68 - 1.06 | 0.149 |
|  |  | Adjusted<sup>a</sup> | 211/1467 | 0.92 | 0.76 - 1.11 | 0.385 | 115/284 | 0.94 | 0.75 - 1.18 | 0.586 |
|  | CVD Mortality | Unadjusted | 34/1467 | 0.86 | 0.56 - 1.33 | 0.492 | 37/284 | 1.17 | 0.79 - 1.72 | 0.442 |
|  |  | Adjusted<sup>a</sup> | 34/1467 | 0.80 | 0.49 - 1.28 | 0.350 | 37/284 | 1.28 | 0.86 - 1.90 | 0.229 |
| **CD34<sup>+</sup>CD133<sup>+</sup> CPCs** | All-Cause Mortality | Unadjusted | 196/1364 | 0.85 | 0.69 - 1.04 | 0.11 | 107/266 | 0.61 | 0.46 - 0.82 | 0.001** |
|  |  | Adjusted<sup>a</sup> | 196/1364 | 0.99 | 0.80 - 1.23 | 0.943 | 107/266 | 0.64 | 0.48 - 0.86 | 0.003** |
|  | CVD Mortality | Unadjusted | 31/1364 | 0.78 | 0.47 - 1.30 | 0.344 | 34/266 | 0.45 | 0.27 - 0.76 | 0.003** |
|  |  | Adjusted<sup>a</sup> | 31/1364 | 0.86 | 0.50 - 1.49 | 0.588 | 34/266 | 0.42 | 0.24 - 0.72 | 0.002** |
| **ECFC** | All-Cause Mortality | Unadjusted | 205/1391 | 0.96 | 0.91 – 1.02 | 0.179 | 116/262 | 0.99 | 0.92 – 1.07 | 0.782 |
|  |  | Adjusted<sup>a</sup> | 205/1391 | 0.97 | 0.92 – 1.02 | 0.261 | 116/262 | 1.01 | 0.93 – 1.09 | 0.828 |
|  | CVD Mortality | Unadjusted | 34/1391 | 1.03 | 0.90 – 1.19 | 0.655 | 37/262 | 0.93 | 0.82 – 1.07 | 0.317 |
|  |  | Adjusted<sup>a</sup> | 34/1391 | 1.03 | 0.90 – 1.19 | 0.633 | 37/262 | 0.95 | 0.82 – 1.09 | 0.437 |

*HR- Hazard Ratio, CI- Confidence Intervals*
*<sup>a</sup>Model adjusted for age, sex, BMI, PAI, Smoking Status, Diabetes Status, Hypertension. \*\*\* P < 0.001, \*\* P < 0.01, \* P < 0.05*

#### Cardiovascular Mortality

Kaplan Meier curves based on tertiles of CPC counts and CV mortality for those free of CVD and those with CVD at exam 8 are shown in **Supplementary Figure 1**. Unadjusted and adjusted Cox proportional hazard models for CPC counts and CV mortality are displayed in **Table 3**. In unadjusted and adjusted Cox proportional hazard models, increases in CD34^+^ and CD34^+^CD133^+^ CPCs were significantly associated with a decrease risk of CV mortality in the CVD present at exam 8 group (all *P <* 0.05, **Table 3**). No other significant associations were observed for CV mortality in either of the sub-groups (all *P* > 0.05).

#### Cardiovascular Events

Cox proportional hazard analysis was performed in the population free of CVD for incidence of future CV events. ECFCs were significantly associated with a decrease risk of future CV events (*P* = 0.046, **Supplementary Table 1**). There was no association between CPC counts and CV event risk for all other measures (all *P* > 0.05).

### Association of Physical Activity with CPC Counts

To assess the association between physical activity and CPC counts, both unadjusted and adjusted linear regressions were performed. In unadjusted and adjusted analyses, PAI and moderate + heavy activity hours were not associated with any CPC subset or with ECFC units. However, in the CVD group, after adjusting for confounders, both PAI and moderate + heavy activity time were positively associated with CD34^+^ CPCs, and were the only significant predictors of the number of these cells (**Table 4** and **Supplementary Table 2**). Physical activity was not associated with CD34^+^CD133^+^, CD34^+^CD133^+^KDR^+^ CPCs or ECFC counts, both in univariate and multivariate analyses.

**Table 4.** Association Between Physical Activity Index and CPC Counts.

| CPC Subset | Outcome | All | CVD-Free | CVD |
| --- | --- | --- | --- | --- |
| | | $\beta$, T-value, P-Value | $\beta$, T-value, P-Value | $\beta$, T-value, P-Value |
| **CD34<sup>+</sup>** | Unadjusted | 0.023, 0.967, 0.334 | -0.013, -0.486, 0.627 | 0.176, 3.009, 0.003** |
|  | Adjusted<sup>a</sup> | 0.008, 0.322, 0.748 | -0.021, -0.813, 0.416 | 0.153, 2.461, 0.014* |
| **CD34<sup>+</sup>CD133<sup>+</sup>** | Unadjusted | -0.014, -0.567, 0.571 | -0.04, -1.495, 0.135 | 0.115, 1.893, 0.059 |
|  | Adjusted<sup>a</sup> | -0.022, -0.872, 0.383 | -0.042, -1.574, 0.116 | 0.107, 1.616, 0.107 |
| **CD34<sup>+</sup>CD133<sup>+</sup>KDR<sup>+</sup>** | Unadjusted | 0.022, 0.901, 0.368 | 0.002, 0.086, 0.932 | 0.108, 1.817, 0.07 |
|  | Adjusted<sup>a</sup> | 0.019, 0.775, 0.439 | 0.005, 0.194, 0.846 | 0.080, 1.269, 0.205 |
| **ECFC** | Unadjusted | 0.005, 0.197, 0.844 | -0.005, -0.201, 0.841 | 0.049, 0.785, 0.433 |
|  | Adjusted<sup>a</sup> | -0.006, -0.241, 0.809 | -0.015, -0.535, 0.592 | 0.036, 0.537, 0.592 |

<sup>a</sup>Model adjusted for age, sex, BMI, Smoking Status, Diabetes Status and Hypertension.
For “All” adjustment also includes previous CVD diagnosis
ECFC- Endothelial colony forming cells.
\*\*\* $P < 0.001$ , \*\* $P < 0.01$ , \* $P < 0.05$

## Discussion

Our main findings were that CD34^+^ and CD34^+^CD133^+^ CPCs were significant predictors of all-cause and CV mortality in the Framingham Offspring cohort, driven primarily by the strength of this association in individuals with CVD. Additionally, increases in self-reported physical activity is positively associated with higher CD34^+^ CPCs in our CVD cohort after adjustment for confounders, a relationship not evident in our CVD free cohort. Together these findings suggest that the observed protection of increased CD34^+^ CPCs on mortality in a diseased population is partly driven by the physical activity levels of individuals.

Several small studies have investigated the prognostic potential of CPCs for predicting incident risk of all-cause and/or CV death. These studies have demonstrated that these cells can predict mortality or clinical end-points in several disease populations, for example patients with coronary artery disease ^18^, acute coronary syndromes ^9^, heart failure ^8^, or type 2 diabetes ^19^. Our data support these observations, with CD34^+^ and CD34^+^CD133^+^ CPCs predictive of all-cause and CV mortality. Interestingly, this association was absent in the CVD-free population, and driven mainly by a strong association with mortality in individuals with pre-diagnosed CVD, suggestive that the prognostic potential of these cells is much stronger in disease populations, and offers little predictive potential, if any, in apparently healthy populations. Interestingly, CD34^+^CD133^+^KDR putative EPCs showed no predictive ability for all-cause or CV death in our study.

In the largest study investigating the role of CPCs on incident risk prediction in a CVD cohort, Patel and colleagues^18^ observed that, like our study, only CD34^+^ and CD34^+^CD133^+^ CPCs were predictive of mortality. In 2 cohorts, each over 400 patients (n=905 pooled), Patel et al ^18^ showed that increases in both these progenitor subsets showed significant inverse association with all-cause and CV-mortality, and that CD34^+^CD133^+^KDR^+^ cells, like our data, showed no association with mortality. Our data in >1700 individuals, however, specifically shows that the associations of CD34^+^ and CD34^+^CD133^+^ with all-cause and CV mortality are driven by their prognostic strength in individuals with CVD, and not those who are CVD-free. It is likely that these cells play a more important role in cardiovascular health when the vascular system is in a state of constant damage, and that lower number of these cells in these patients reflect exhaustion of the progenitor cell pool.

Both CD34^+^ and CD133^+^ progenitor cells have vascular regenerative capabilities ^20^. These cells, reported initially to have pro-angiogenic capabilities due to the potential to differentiate into endothelial cells ^1^, most probably work in a paracrine manner, through secretion of vasoactive and proangiogenic factors, such as VEGF and other pro-angiogenic cytokines ^21^. Due to their potential vasculo-reparative capacities, clinical studies have been undertaken to assess their efficacy as cellular therapies to promote recovery of blood flow in myocardial infarction and stroke studies. Clinical studies showing implantation or injection of these cell types show promise in repair of damaged myocardium in animal models ^2^ and in some human studies ^22, 23^, however due to the expense and research and development required to optimize this cellular therapy, other non-pharmaceutical interventions may be more effective in promoting endogenous vascular repair for clinical benefit. In addition given the reduced number and function of these cells in CVD ^5^, and the predictive association with mortality, it is pertinent to find therapies to augment production, mobilization and function of these cells. Exercise and physical activity has the potential to mobilize these cells into the circulation as evident from acute exercise studies showing transient increases in CPCs in both healthy^11, 24^ and diseased populations ^25^, although the response to acute exercise is somewhat diminished in CVD patients ^26^. Long-term physical exercise and physical activity shows promise in increasing number and/or function of these CPCs ^27^, potentially through promoting bone marrow production of progenitor cell subsets (although the origin of EPCs has been a topic of debate recently^28^) or via reducing inflammatory or pro-apoptotic stimuli in the circulation ^29^, thus enhancing survival of these cells in our body. Our data support the use of physical activity to promote or maintain CD34^+^ CPC number in humans. High levels of self-reported physical activity was associated with reduce risk of all-cause and CV mortality (data not shown), and interestingly was associated with higher number of CD34^+^ and CD34^+^CD133^+^ CPCs which were also associated with mortality, but only in individuals with CVD, and not in our CVD-free group.

Together these findings suggest that the observed protection of increased CD34^+^ CPCs on mortality in a diseased population is partly driven by the physical activity levels of individuals. These findings maybe clinically relevant as it is supportive of exercise-based cardiac rehabilitation and suggests an area for future interventions. Whilst both acute aerobic ^24^ and resistance exercise ^11^ can promote progenitor cell release and improve pro-angiogenic function, long-term resistance exercise training studies are lacking and thus warranted, specifically in a CVD cohort.

## Conclusion

Our study demonstrated that CD34^+^ and CD34+CD133^+^ CPCs are predictive of mortality in a large cohort, but with more prognostic potential in individuals with CVD. Additionally, physical activity was associated with significantly greater CD34^+^ CPCs in a CVD population, with no relationship in non-CVD population. Exercise and physical activity may promote vascular health and longevity in CVD patients via promoting CD34^+^ CPC number.

## Data Availability

The data underlying this article were provided by the National Heart, Lung and Blood Institute (NHLBI) under license / by permission. Data will be shared on request to the corresponding author with permission of the NHLBI.

## Acknowledgements

This manuscript was prepared manuscript was prepared using FRAMOFFSPRING Research Materials obtained from the NHLBI Biologic Specimen and Data Repository Information Coordinating Center and does not necessarily reflect the opinions or views of the FRAMCOHORT, FRAMOFFSPRING or the NHLBI.

## Authors Contributions

M.R conceived and designed the research. D.M, M.R and J.D undertook statistical analysis of the data. M.R and D.M interpreted results of the experiments, prepared figures and drafted the manuscript; all authors edited and revised the manuscript; all authors approved the final version of the manuscript.

## Supplementary information

### Data analysis

Data analyses were carried out using RStudio Team (2019, RStudio Team (2019). RStudio: Integrated Development for R. RStudio, Inc., Boston, MA: http://www.rstudio.com/) utilising the R core package [1] and the extension packags “dplyr” for data clean ing and transformation [2], “survival” for survival analyses [3] and “ggplot2” [4].and “survminer” [5] for figure generation.

## Supplementary Tables and Figures

**Supplementary Table 1.** CPC Counts and risk of future CV events in participants free of CVD diagnosis at exam 8.

| CPC Subset | Outcome | Model | No. of<br>events/ No.<br>at risk | HR | HR 95%<br>CI | P value |
| --- | --- | --- | --- | --- | --- | --- |
| CD34<sup>+</sup> CPCs | CVD Event | Unadjusted | 191/1467 | 0.99 | 0.75 - 1.30 | 0.925 |
|  |  | Adjusted<sup>b</sup> | 191/1467 | 1.18 | 0.88 - 1.59 | 0.279 |
| CD34<sup>+</sup>CD133<sup>+</sup>KDR<sup>+</sup> EPCs | CVD Event | Unadjusted | 191/1467 | 1.09 | 0.91 - 1.31 | 0.372 |
|  |  | Adjusted<sup>b</sup> | 191/1467 | 1.06 | 0.87 - 1.28 | 0.588 |
| CD34<sup>+</sup>CD133<sup>+</sup> CPCs | CVD Event | Unadjusted | 177/1364 | 0.97 | 0.77 - 1.21 | 0.783 |
|  |  | Adjusted<sup>b</sup> | 177/1364 | 1.05 | 0.83 - 1.33 | 0.688 |
| ECFC | CVD Event | Unadjusted | 187/1391 | 0.94 | 0.89 - 1.00 | 0.052 |
|  |  | Adjusted<sup>a</sup> | 187/1391 | 0.94 | 0.88 - 1.00 | 0.046* |

HR- Hazard Ratio, CI- Confidence Intervals.
<sup>a</sup>Model adjusted for age, sex, BMI, PAI, Smoking Status, Diabetes Status, Hypertension
\*\*\* $P < 0.001$ , \*\* $P < 0.01$ , \* $P < 0.05$

**Supplementary Table 2.**
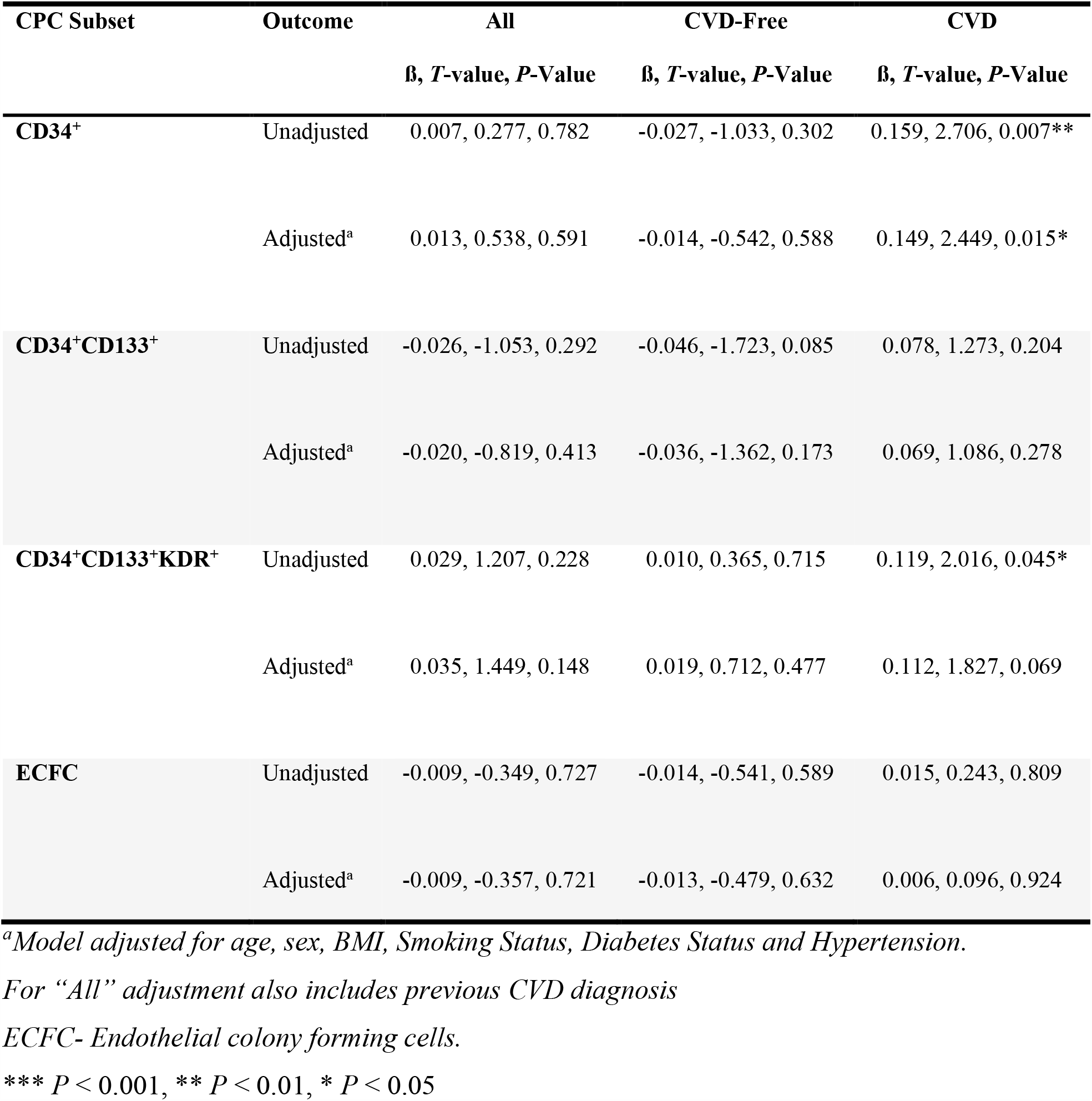
Association Between Time Spent at Moderate+Heavy Physical Activity and CPC Counts.

**Supplementary Figure 1.**
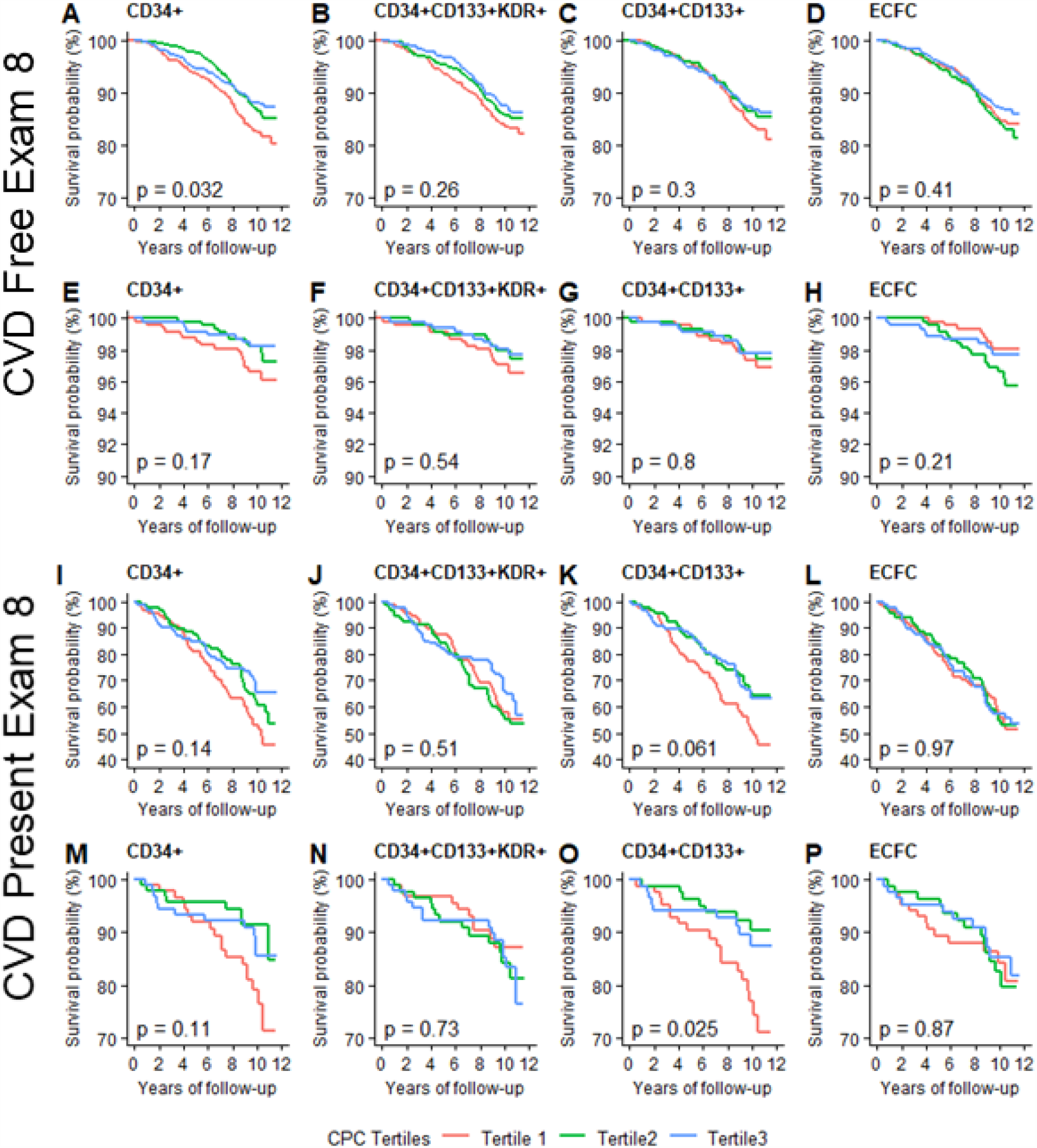
Kaplan Meier Survival Curve’s for the relationship between mortality and CPC tertile split by participants free of CVD at exam 8 (A – H) and those diagnosed with CVD at exam 8 (I – P). Curves A-D and I – L are for all-cause mortality and curves E – H and M - P are for CV mortality. (Tertile 1 = Low count, Tertile 2 = Moderate count, Tertile 3 = High count). Statistical significance was set at p <0.05.

